# How did Ontario healthcare institutions implement and legitimize Covid-19 vaccine mandates? A mixed-methods study protocol

**DOI:** 10.1101/2025.05.11.25327408

**Authors:** Claudia Chaufan

## Abstract

**Background:** Upon the WHO declaration of COVID-19 as a pandemic, healthcare workers (HCWs) - unvaccinated by necessity - were celebrated as “heroes” for their service under difficult conditions. Later, as some of them resisted vaccine mandates, they were reframed as “threats”, regardless of personal behaviour, workplace setting, or evidence of their harmfulness. This discursive shift and the institutional mechanisms supporting it remain underexamined.

**Goal:** This protocol outlines a mixed-method study investigating the implementation of COVID-19 vaccine mandates in medical establishments across Ontario, Canada.

**Methods:** This is a two-phase mixed methods planned study. Phase 1 is an environmental scan of institutional vaccine mandate policies across a purposive sample of diverse Ontario medical establishments. It will track policy implementation timelines, mandates scope, exemptions eligibility criteria, and supporting scientific evidence presented. Phase 2 is a critical interpretive analysis of documents collected in Phase 1 that draws on Max Weber’s theory of bureaucracy and legitimacy, Carol Bacchi’s “What Is the Problem Represented to Be” (WPR) approach, and Brian Martin’s framework on suppression of dissent to identify patters in the institutional framing and treatment of challenges to mandated vaccination.

**Expected Outcomes:** This study is expected to yield descriptive and interpretive insights into how bureaucratic structures shaped mandate enforcement and dissent suppression. Results are expected to inform academic debates on institutional legitimacy, governance, and public health ethics.

## Introduction

This study is motivated by a paradox that began to unfold in the early months of the COVID-19 policy response: healthcare workers (HCWs), unvaccinated by necessity, were celebrated as heroes for their willingness to serve under crisis conditions (see for example (Simmons, 2020a, 2020b)). Yet many of these same workers would be later reframed as threats to public health - regardless of their protective behaviours, workplace settings, or evidence of their harmfulness (see for example (Castanet News, 2021; D’Avino, 2024)). This shift - from “hero” to “threat” - raises critical scientific, policy, and ethical questions. Why were vaccine mandates implemented as conditions of employment for those already exposed to high levels of risk? What evidence and arguments were presented to support the contention that vaccination would prevent viral transmission in healthcare settings, the key rationale for their implementation? How did political and institutional dynamics shape the justifications and enforcement of mandates? And how were dissent, resistance, or requests for accommodation framed within policy discourse?

This protocol outlines a qualitative and policy analytic study that will investigate the implementation of COVID-19 vaccine mandates in medical establishments in Ontario, Canada. While many studies have focused on what has been framed as “vaccine hesitancy”, we focus instead on mandate legitimation and dissent suppression as institutional processes. This examination will document the timeline of mandate introductions, appraise how institutions interpreted and operationalized provincial directives, and identify what scientific, administrative, or rhetorical justifications were used to support their adoption. The work will be grounded in a critical analysis of the social and institutional narratives that shaped HCWs’ changing roles throughout the COVID policy response. Instances of such narratives appeared following the policy rollout when the “hero” label was deployed as a technique of governance and a tool to enforce professional compliance. As Mohammed et al have argued, the language of heroism - replete with militaristic and sacrificial imagery - elevated frontline HCWs while masking serious lapses in workplace safety, labour protections, and staffing shortages (Mohammed et al., 2021). Similarly, Boulton et al. have suggested that public rituals such as balcony clapping and military flyovers further served as forms of “performative allyship” that, while seemingly supportive, helped to normalize institutional neglect by recasting structural vulnerability as moral duty (Boulton et al., 2022). These worthy critiques notwithstanding, to our knowledge no research has been conducted on the rhetorical and institutional reframing of dissenting HCWs from lauded protectors of public health to perceived threats to it, thus our planned study.

Indeed, since the launch of the global vaccination campaign, much of the literature that addresses HCWs’ concerns with or refusal of vaccination has tended to pathologize dissent, focusing on behavioural correction through education or coercion rather than engaging with the scientific, political, or ethical dimensions of resistance (see for example (Casey et al., 2022; Emanuel & Skorton, 2021)). Rarely considered are the moral and psychological tolls that these policies have imposed - particularly on women, who already face higher rates of burnout, moral distress, and suicide (K. A. Lee & Friese, 2021; Peters, 2025). Finally, even less explored is the institutional silence surrounding the demonization of dissenting HCWs, some of whom have faced termination, ridiculing, or blacklisting – or have even witnessed hostility towards unvaccinated patients (Chaufan et al., 2025b). These gaps in the literature suggest not only a lack of scholarly curiosity but also a tendency to frame resistance as pathology rather than as a potentially legitimate professional - scientific and ethical - stance.

The study builds on a body of work by the lead author and collaborators, including critical analyses in the academic literature of problem representations of HCWs’ dissent (Chaufan & Hemsing, 2024), surveys on the impacts of COVID-19 policy on HCWs’ well-being and patient care (Chaufan et al., 2024, 2025a), thematic analyses of HCWs lived experience (Chaufan et al., 2025b), and the medicalization of dissent with official COVID policies (Chaufan, 2023, 2024) . It is part of two larger projects registered in the Open Science Framework that examine the impact of COVID-19 policies on healthcare work and governance more broadly (https://osf.io/z5tkp ; https://osf.io/84kbr/). By examining the tensions in the healthcare workplace through the lens of policy implementation, institutional authority, and discourse, the study aims to illuminate a neglected dimension of the COVID-19 policy response: how policies framed as protective may have functioned as instruments of exclusion, control, or compliance enforcement - especially against those within the healthcare system who raised difficult, but necessary, questions.

### Theoretical Framework

The interpretive phase of this study is grounded in the sociology of Max Weber, particularly his conceptualizations of bureaucracy, rational-legal authority, and ideal types. These concepts will help to illuminate how healthcare institutions implemented COVID-19 vaccine mandates, legitimized those decisions, and framed or managed internal dissent within their bureaucratic structures. Weber’s model of bureaucracy emphasizes rule-governed conduct, hierarchical organization, and a clear distinction between personal and official roles. These features were especially pronounced at the launch of the global vaccination campaign, as healthcare institutions invoked procedural authority to justify exclusionary mandates. That said, and as Byrkjeflot has argued (Byrkjeflot, 2018), Weber’s model has often been misinterpreted as primarily about efficiency and rule-governed behaviour in the context of hierarchical institutions, when it could be as much or better understood as a theory of legitimacy - an insight central to this study’s analysis of institutional behavior.

To organize institutional variation in mandate implementation, the study will also apply Weber’s concept of “ideal type” (Swedberg, 2018), an analytical construct that emphasize certain features of a phenomenon in order to enable comparison. Ideal types will be used to develop heuristic categories of institutional response, ranging from minimalist compliance with provincial guidance to maximalist exclusion of unvaccinated staff. These categories are not meant to capture reality perfectly, but to provide clarity in identifying patterns. As well, this analysis draws on Weber’s notion of “verstehen,” or “interpretive understanding”. As Gann explains (Gann, 2017), verstehen is about grasping the subjective meaning of action. Our project will apply this interpretive lens to institutional texts - mandate statements, internal memos, media releases - in order to identify not only what was *said*, but also what was *meant* – therefore supplementing the environmental scan with a richer understanding of how mandates were discursively justified to render dissent invisible or illegitimate.

Finally, the application of Weberian concepts - bureaucracy, ideal types, and verstehen – framing the second phase of the project will be informed by Brian Martin’s typology of dissent suppression (Martin, 1999), which will help to document how dissenting HCWs were not only excluded in practice, but also delegitimized through tactics such as devaluation, reinterpretation, and institutional silence. This typology will allow the study to go beyond documenting policy timelines and examine how authority, legitimacy, and exclusion were constructed in real time. By analyzing the treatment of dissenting HCWs through the lens of this typology, the project aims to document whether and how such forms of suppression operated in institutional settings during COVID.

**Table 1.**
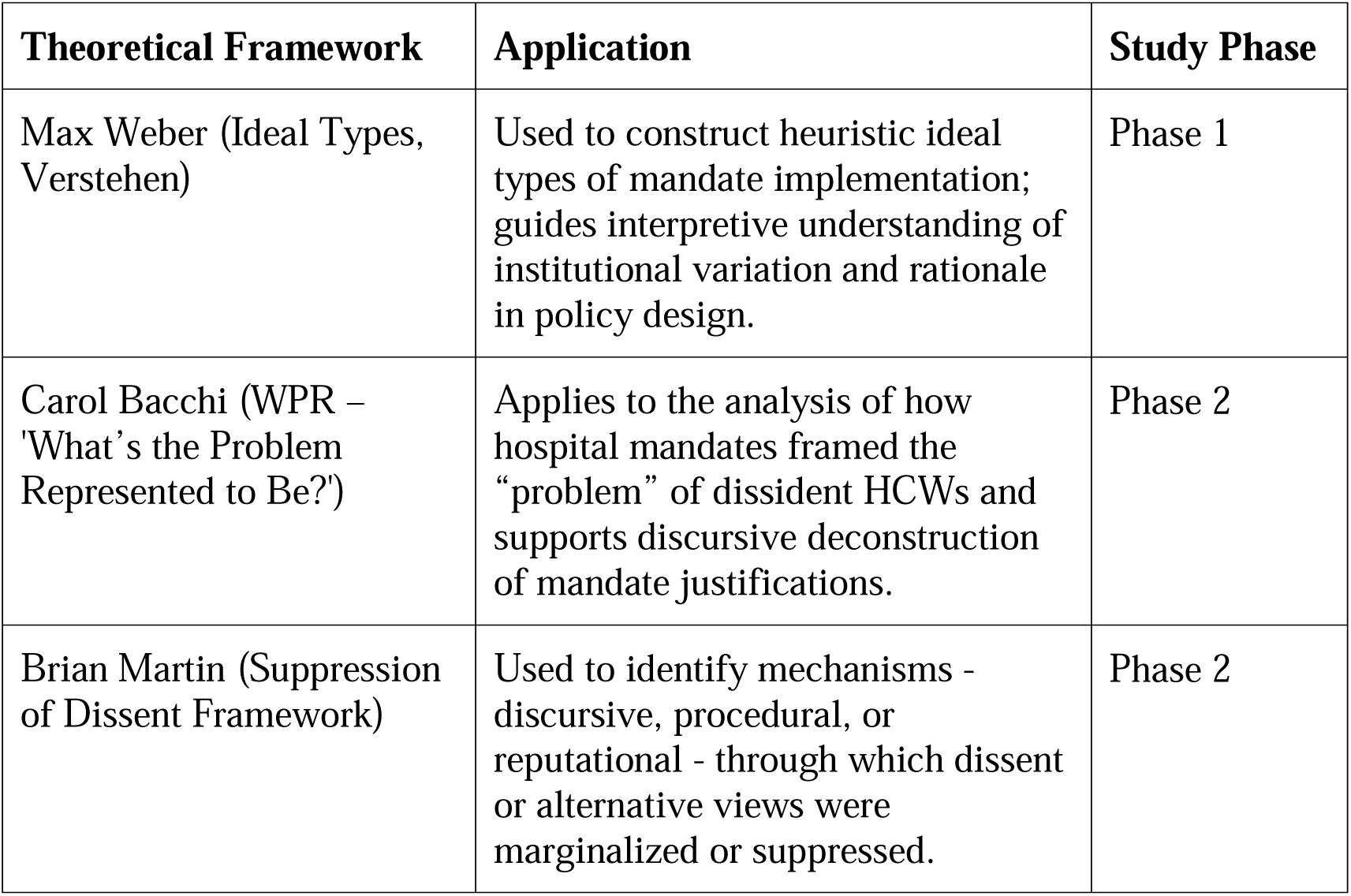
Mapping theoretical frameworks to study phases. This table presents the alignment between the study’s theoretical frameworks and the two main phases of research. Each framework contributes a distinct analytical perspective to guide data collection, categorization, and interpretation.

### Aim and objectives

The overall aim of this study is to document the implementation of COVID-19 vaccine mandates in medical establishments in Ontario, Canada, understand how authority, legitimacy, and exclusion were constructed in real time, and elaborate on the scientific, political and ethical implications of these processes. The following research questions will guide the study: 1) How and when were COVID-19 vaccine mandates implemented across medical institutions in Ontario? 2) What were the institutional rationales, and how were alternatives framed, dismissed, or suppressed?

### Design

The design consists of two sequential and interrelated parts: (1) an environmental scan of institutional vaccine mandate policies in Ontario medical establishments, and (2) a critical interpretive analysis of these policies using document analysis and the theoretical lenses and tools described earlier. Below we describe the two phases:

- Phase 1: this *descriptive* phase will involve an environmental scan, a method developed for the business sector (Choo, 2001) yet also used in other sectors - such as education (Pashiardis, 1996) and increasingly health (Charlton et al., 2021) - to identify and document policy formulation, development and implementation patterns across institutions. The scan will focus on Ontario medical establishments and publicly-available documents will be extracted from institutional websites, media releases, internal memoranda when accessible, and communications from provincial public health authorities - especially Directive #6 issued by Ontario’s Chief Medical Officer of Health (Ontario Health, 2021). A structured table will track: 1) the timeline of mandate implementation, duration, reversals or modifications; 2) alternatives to vaccination permitted; 3) references to scientific justifications; and 4) potential externals influences and responses to policies in the health sector (e.g., media articles; press releases).
- *Phase 2*: this *interpretive* phase will apply document analysis and critical policy analysis to examine how institutions framed their decisions and suppressed dissent. This analysis will draw from Carol Bacchi’s WPR framework, which asks what “problem” a policy presupposes to solve, and how such “problem representations” structure the boundaries of available responses. The study will also draw on Brian Martin’s typology of the suppression of dissent in science, which includes strategies such as devaluation, reinterpretation, intimidation, and reliance on official channels. These frameworks will guide the examination of how dissenting HCWs were portrayed and managed in institutional discourse, particularly where mandates were framed as unassailable “evidence-based” imperatives.

Each phase will contribute differently but complementarily to answering the overarching research questions, as they will jointly generate both a *descriptive* mapping of vaccine mandate policy trajectories and a *theoretically informed analysis* of institutional discourse. This dual structure aims to produce a layered understanding of how mandates were justified, operationalized, and contested in the Ontario healthcare sector. The findings are also expected to inform broader discussions about the use of public health authority in institutional decision-making and its implications for professional autonomy, scientific debate, and democratic accountability.

### Data

We will select hospitals for analysis using data extracted from the *Canadian Broadcasting Corporation [CBC]’s Rate My Hospital* database, which reports having evaluated approximately 240 acute-care hospitals across Canada, including 153 in Ontario (CBC News, 2024). Each Ontario hospital was categorized by the CBC according to two criteria: (1) type of institution (Teaching, Large Community, Medium Community, or Small Community) and (2) performance grade, based on a weighted composition of outcome and quality indicators. Grades ranged from A+ (highest) to A, B, C, and Unrated (for hospitals with insufficient data to assess performance).

To ensure that our qualitative policy analysis captures the full diversity of institutional configurations and policy responses, we have constructed a 20-cell matrix that intersects hospital type with grade. We will place all CBC ranked Ontario hospitals into their respective matrix cells and select a purposeful sample by drawing at least one hospital from each populated cell, a “maximum variation sampling” approached used to document “unique or diverse variations that have emerged in adapting to different conditions” and to identify salient patterns across variations (Palinkas et al., 2015)(p.3). By sampling internal documents from medical establishments across all filled cells, we aim to balance representational breadth with feasibility. We anticipate selecting a sample of 20 - 25 institutions for in-depth analysis. Press releases issued by hospitals and media reports of hospital policy will also be included in the data set to strengthen the diversity of sources accounting for institutional behaviours.

### Expected Outcomes

This study will yield descriptive and interpretive insights into how bureaucratic structures shaped mandate enforcement and dissent suppression. Results should inform academic debates on institutional legitimacy, governance, and public health ethics through a variety of strategies of dissemination. In practice, they will culminate in 4 publications:

1. A descriptive report that maps mandate implementation and variation across institutions using environmental scan methodology.
2. A critical analysis of the rationale and discourse surrounding the mandates, drawing on Carol Bacchi’s “What is the Problem Represented to Be?” (WPR) framework (Bacchi, 2016), Martin’s typology, critical discourse analysis (Fairclough, 2013), thematic analysis (Braun & Clarke, 2006) and document analysis (Bowen, 2009).
3. A policy brief that integrates and translates the results of the study for a policy audience.
4. A lay article that integrates and translates the results of the study for the public.

### Statement on Reflexivity

This study is informed by the lead investigator’s long-standing professional interest in the intersection of policy, governance, and the ethics of healthcare delivery. While the author holds well-developed views about the political and institutional dynamics surrounding COVID-19 mandates, the research design is grounded in a commitment to methodological rigour and epistemic integrity. The aim is not to confirm predetermined assumptions but to systematically document and interpret the evidentiary and discursive basis of institutional decision-making.

Following Finlay (Finlay, 2002), reflexivity is approached here not as a means of personal disclosure but as a discipline of methodological transparency and interpretive accountability - meaning not foregrounding the researchers’ biographies but rather as reflexive practice deployed to clarify how judgments were made about source selection, interpretive framing, and categorization of mandates. This reflexive orientation allows research like this study to maintain a dual commitment, both to rigorous analysis of institutional policy and to critical self-awareness about how the researcher’s position may shape - not predetermine - interpretation.

### Limitations

It may be argued that this project has a built-in bias due to his negative framing of mandated vaccination and is therefore incapable of informing policy. To this we reply that while we have our position on the policy of mandates, the study will not presume their legitimacy or lack thereof, but rather document and explain how they were implemented, justified and contested, and what the tensions identified may reveal about institutional decision-making and professional agency. Critics may also question whether we can expect to gather enough data to support our theoretically informed analysis; they may also question whether the CBC ranking, while offering a consistent starting point for institutional sampling, can truly capture policy diversity or institutional discretion, given the shortcomings of its evaluation criteria, as acknowledged by the CBC itself. We believe that despite these shortcomings – shared by comparable studies, as authors acknowledge (Curran et al., 2024) - the interpretive phase will be grounded in a diverse and robust corpus of texts – institutional documents and media reports – that should ensure that theoretical insights are empirically anchored, enough to make this study valuable and transferable to other settings and jurisdictions (Drisko, 2025).

### Significance

This study should offer timely and broadly relevant insights into how healthcare institutions implemented and justified COVID-19 vaccine mandates, while offering a novel perspective, by treating vaccine mandates not merely as clinical interventions, but as institutional policies subject to political, rhetorical, and ethical contestation. By focusing on Ontario - Canada’s largest province and a leader in healthcare policy - the study will provide a detailed case that can serve as a framework for similar investigations across other provinces. Given that healthcare in Canada is primarily administered and delivered at the provincial level - with federal guidance providing general standards but not operational control (Government of Canada, 2016) - understanding institutional decision-making at the provincial level is essential for evaluating the health system under contested conditions.

The project is also significant for what it can reveal about institutional behaviour in the face of political pressures. While prevailing narratives framed the moment as an emergency (Gray & Stone, 2020), responses to vaccine mandates varied widely - not only across jurisdictions, but also between institutions operating under the same provincial directives. These variations raise important questions about how formal organizations enact authority, interpret policy, and manage internal dissent. Max Weber’s analytical lens - particularly his insights into bureaucratic rationality and the interpretive meaning of social action – will help to examine how institutions balanced the tensions between competing goals – providing high quality patient care, complying with public health directives, managing internal / sectoral politics, and maintaining internal and external legitimacy. It will also help to understand the tension between structure and agency, i.e., the discretionary space available to decision-makers within bureaucratic systems and the details of the use of this space by medical institutions under the perceived weight of provincial orders.

In addition to these institutional and professional implications, the study may also have relevance for patient outcomes and health care quality. By exploring how healthcare institutions implemented and justified mandates, the research can help illuminate how administrative decisions - shaped by legal authority, rhetorical framing, and organizational discretion - may influence staffing levels, workplace morale, and institutional trust, which in turn can affect the quality, continuity, and equity of care delivered within healthcare systems (Amiri et al., 2019; Austin et al., 2017), especially under conditions of institutional uncertainty and operational strain.

Further, the project should also contribute to broader debates about how legitimacy is constructed and enforced in public health governance, by documenting not just policy timelines but also rhetorical strategies, decision-making logics, and institutional responses to dissent. In doing so, it should shed light on questions of professional autonomy, evidence-based policy, and the ethics of labour practices in the health sector. Moreover, findings can shed light beyond Canada. For example, in the United Kingdom, with a more centralized health care policy, the government initially announced plans to mandate Covid-19 vaccination for National Health Service workers (J. Lee & Jackson, 2021), but in contrast to Canada, it reversed the policy partly due to organized resistance from within the profession (Gregory, 2022). In the United States, where health policy is more decentralized than in Canada, some states – for instance, California - enforced healthcare mandates (Feder Ostrove, 2021), while others - Texas and Florida - banned them altogether (Roth, 2021). These divergent approaches highlight the need for grounded, context-sensitive analysis. By documenting the Ontario case in detail, this project should offer conceptual and empirical tools to examine how institutions across jurisdictions justify policies involving coercion, manage opposition, and maintain legitimacy in the name of public health.

### Ethics

This study relies solely on publicly available data so does not require ethics review.

## Author Contributions (CRediT taxonomy)

Conceptualization: Claudia Chaufan

Methodology: Claudia Chaufan

Writing – Original Draft: Claudia Chaufan

Writing – Review & Editing: Claudia Chaufan

Investigation (anticipated): Natalie Hemsing, Maryanne Dias

Formal Analysis (anticipated): Natalie Hemsing, Maryanne Dias

Project Administration: Claudia Chaufan

*Note:* NH and MD are expected to contribute to subsequent phases of the project, including data collection and interpretive analysis. Authorship on future publications will reflect the nature and extent of each contributor’s involvement in accordance with ICMJE guidelines.

## Data Availability

All data in the present protocol are contained in the manuscript

